# Virus Genome Sequences in the Blood of Myalgic Encephalomyelitis/Chronic Fatigue Syndrome Patients

**DOI:** 10.1101/2025.11.06.25339689

**Authors:** Richard W. Hyman, Weihong Xu, Kevin R. Roy, Sundari Suresh, Robert P. St.Onge, Ronald W. Davis

**Affiliations:** Department of Biochemistry, Stanford University School of Medicine, Stanford, CA; Stanford Genome Technology Center, Stanford University, Palo Alto, CA; Greater Bay Area Institute of Precision Medicine (Guangzhou), Fudan University, Guangzhou, China; Department of Genetics, Stanford University School of Medicine.

## Abstract

Myalgic Encephalomyelitis/Chronic Fatigue Syndrome (ME/CFS) is a baffling disease. The disease has a wide spectrum of severity, to date has no established molecular marker, no known causation, and no cure. Many patients report in retrospect that they suffered a virus infection prior to suffering their first symptoms of ME/CFS. Therefore, we report a search for virus genome sequences in the cell-free blood of ME/CFS patients and healthy controls. We used a panel of molecular probes to assess the presence or absence of 185 diverse human viruses in each sample. We identified a total of seventeen viruses, with more in the healthy controls than in the ME/CFS patients.

## Introduction

ME/CFS is reported to affect over one million patients in the USA alone (Committee on the Diagnostic Criteria for Myalgic Encephalomyelitis/Chronic Fatigue Syndrome et al. 2015; Cortes Rivera et al. 2019; Valdez et al. 2019). While the symptoms vary widely, ME/CFS can be very debilitating. The diagnosis of ME/CFS relies, in part, on ruling out other known causes (Bateman et al. 2021). Since many patients report in retrospect that they suffered a virus infection prior to the onset of ME/CFS (Rasa et al. 2018), it is possible that ME/CFS is the result, at least in part, of a chronic virus infection. We have employed molecular probe technology to detect virus genome sequences in the cell-free blood of ME/CFS patients and healthy controls. We identified a total of seventeen viruses, with more in the healthy controls than in the ME/CFS patients.

## Materials and Methods

### 1. Participants

All participants gave informed consent. The drawing of blood for these studies was approved by the Stanford University IRB (Protocol 34830). Details of the participants have been published (Chang et al. 2021). ME/CFS patients are designated by numbers below 100. Healthy controls were numbered starting with 100.

### 2. Nucleic acid isolation

Cell-free nucleic acids were prepared employing the QIAamp Circulating Nucleic Acid Kit (Qiagen) as described by the manufacturer. The nucleic acids were dialyzed and concentrated by use of a Centricon (Millipore). The nucleic acids were subjected to whole-genome amplification and reamplification by employing GenomePlex Complete Whole Genome Amplification (and Reamplification) Kits from Sigma-Aldrich. For each DNA, multiple reamplification reactions (often 48) were run in parallel. The products were combined, dialyzed, and concentrated in a Centricon.

### 3. Homers

The essence of our molecular probe method is the identification of 60 base sequences unique to each virus genome. We call these “Homers”. The Homers were derived from the genome sequences available on the National Center for Biotechnology Information public website (RefSeq) (https://www.ncbi.nlm.nih.gov/genome/browse/#!/overview/) by applying the custom script, *blaster.rb*, which is archived with Zenodo at https://doi.org/10.5281/zenodo.17391545. In essence, starting at one end of the sequence 60 consecutive bases were selected. These were required to be between 40% and 60% adenine-plus-thymine. No base was allowed to appear more than three times in a row. 60-base sequences passing those screens were blasted against GenBank. Those 60-base sequences unique to that virus genome were identified and selected. We call these “Homers”. Then, the selection process was moved 30 bases on the genome sequence and the process repeated, and so forth.

### 4. Molecular probe design

The design of the molecular probes has been published (Hyman et al. 2012; Xu et al. 2014). In brief, the 60-base Homers were divided into two 30-base sequences separated by a universal 36-base sequence yielding a 96-base oligonucleotide (Figure 1A). Thus, the identical 20-base forward and reverse amplification primers, which overlap four bases at their 5’ ends, amplified all probes. The last step in probe design was to enter the sequence into the Website DNA Folding Form (https://www.unafold.org/mfold/applications/dna-folding-form.php) (Zuker 2003). Any design that produced a form that included eight or more base pairs was discarded.

**Figure 1.**
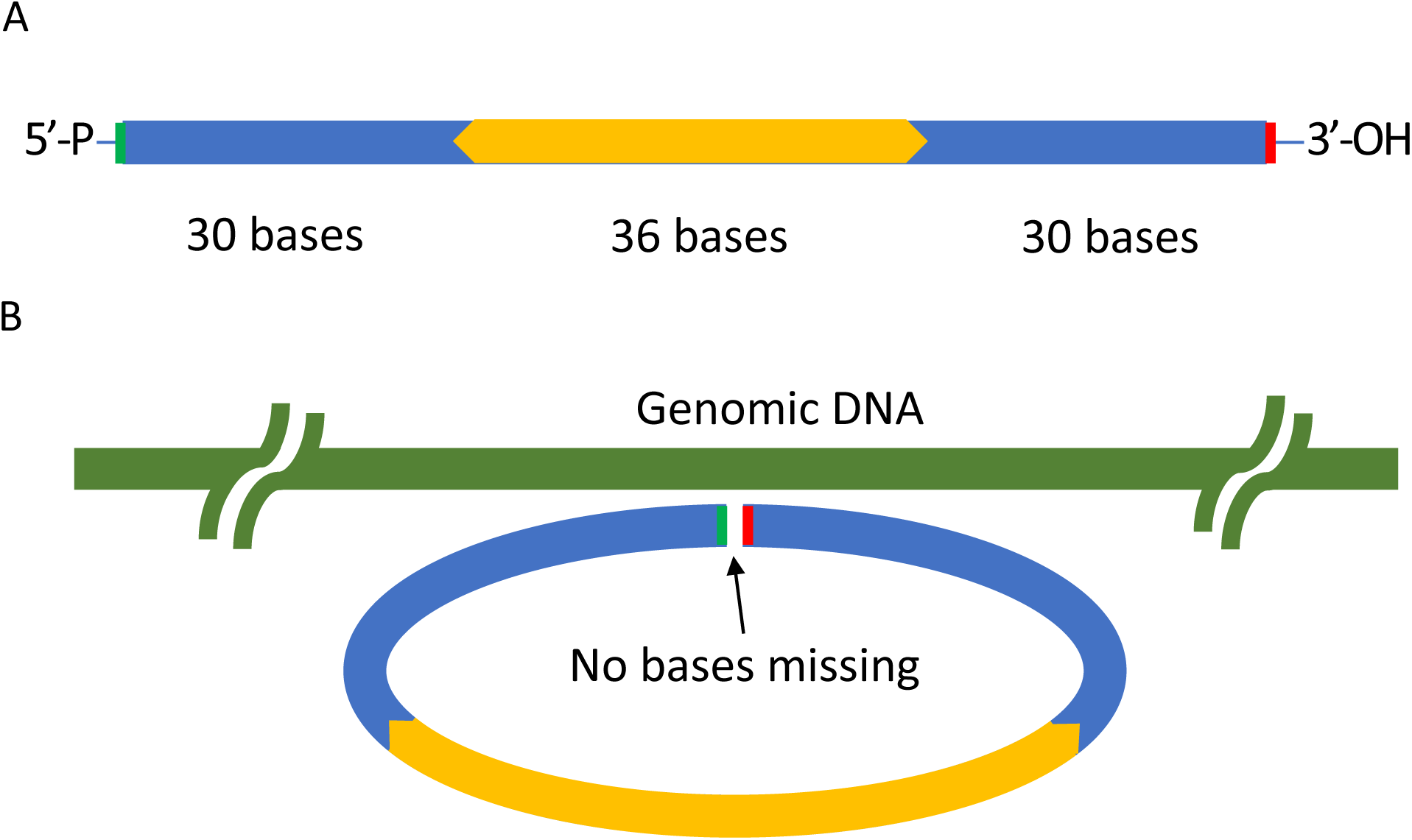
Molecular probe technology. (A) Molecular probe design. The 60-base Homer is split into two end segments of 30 bases each. The central section of 36 bases is common to all probes and is complementary to the 20-mer forward and reverse primers that overlap by four bases at their respective 5’ ends. The green color on the left end represents a 5’ phosphate group. The red color on the right end represents a 3’ hydroxyl group. (B) Probe hybridized to its target. When a probe is hybridized to its target, the 5’-phosphate is adjacent to the 3’-hydroxyl group. No bases are missing. Therefore, the enzyme DNA ligase will synthesize a phosphodiester bond to create a circular, single-stranded DNA.

### 5. Molecular probe ensemble

The molecular probe ensemble was composed of 1508 oligonucleotide probes representing 185 virus genome sequences (Figure S1, Supplementary Information). Thus, the average was 8.2 probes per virus genome. The range was wide: from a high of 40 probes for West Nile virus, and, at the low end, only one probe per virus for 26 viruses. For a total of 140 virus genomes, there were, at least, three probes per genome. The molecular probe ensemble was purchased from Agilent Technologies and processed as previously described(Krishnakumar et al. 2008; Xu et al. 2014; Smith et al. 2017). PCR amplification was employed to produce a working stock of the molecular probe ensemble. Following PCR amplification, the molecular probe ensemble was converted to single-stranded DNA as previously described (Krishnakumar et al. 2008; Xu et al. 2014). In the final step of probe production, the 5’ ends of the single-stranded DNA probes were phosphorylated by T4 polynucleotide kinase.

When the 96-base probe has hybridized to its homologous target sequence, the 5’-phosphate was adjacent to the 3’-hydroxyl group (Figure 1B) with no gap in between. Importantly, no bases were missing. The enzyme DNA ligase was employed to ligate the 96-base oligonucleotide into a covalently closed circle. After removal of unreacted probes by exonuclease digestion and clean-up, PCR was employed with the universal forward and reverse primers to amplify the sequence of interest. Barcodes were included in the amplification primers just upstream of the 20-base forward and reverse probe priming sites so that they would be read out at the beginning of read 1 and read 2, respectively (Table S1, Supplementary information). A unique barcode combination was used for each sample (Table S2, Supplementary information). The amplified products were sequenced by Novogene USA on the Illumina HiSeq 2500 (20191112 ME/CFS patient samples) and by the Stanford Genome Technology Center on the Illumina NextSeq 550 (20200804 healthy control samples), all with paired-end 150 bp short read sequencing.

### 6. Sequence read analysis

All scripts, keyfiles, and data (starting with the collapsed fastq sequences at the end of step2) are available at https://github.com/k-roy/MOLECULAR_PROBES and archived with Zenodo at https://doi.org/10.5281/zenodo.17429326. Supplementary Tables S1-S3, including the sequencing primers, primer mixes, and probe counts are available below and as an Excel file in the Zenodo repository. The sequence read processing steps are briefly summarized below.

Step 1. The reads belonging to the molecular probe amplicons were retrieved from the unindexed read files from each sequencing run by searching for 11 b unique sequences corresponding to the forward (5’-AGGTCGTACA-3’) and reverse (5’-GTGACTATCGA-3’) priming sites in read 1 and read 2, respectively.

Step 2. The amplicons are 112 bp, consisting of the probe sequences, universal priming sites, and the 6-mer barcodes, such that both read 1 and read 2 completely spanned the inserts. To combine the reads, we first trimmed Illumina TruSeq adapters from the 3’ ends with bbduk using the kmer 5’-AGATCGGAAGAGC-3’ (Bushnell et al., 2014), then merged read 1 and read 2 into a single synthetic read with bbmerge (Bushnell et al. 2017), followed by collapsing identical reads and inserting the read count into each read name.

Step 3. The 60 bp Homer sequences were extracted from each merged and collapsed read. Firstly, the sequences were searched for perfect matches against the 1,439 designed probes. To account for sequencing errors, partial matches were also allowed where either the first 20 bp or last 20 bp of the designed probe was found in the read. The perfect and partial match counts were summed for each sample to generate a table with probe IDs as rows and samples as columns with the probe counts in each cell (Table S3, Supplementary information).

### 7. Differential analysis

DESeq2 was first applied to identify molecular probes enriched in ME/CFS patients relative to negative controls (Love et al., 2014). Probe-level log2 fold changes and standard errors were then aggregated to the virus level using inverse-variance weighting, yielding pooled fold changes and corresponding errors. Virus-level p-values were computed employing the Simes combined p-value as this method is more robust to zero counts (Ham and Park 2022). Statistical significance was defined as log2-fold change >1 and p-value <0.05 for both molecular probes and virus throughout this manuscript. These bioinformatics analyses were performed in R (version 4.4.0). The volcano plot (Figure S2, Supplementary information) was generated with *ggplot2*, and the heatmap with pheatmap.

## Results

The data are expressed as the number of high-throughput sequencing reads attracted to each molecular probe for each clinical sample. These are collected in Table S3 (Supplementary information). To save space, only those probes that attracted at least ten sequence reads in at least one clinical sample are included in the table. DESeq2 (Love et al. 2014) was applied to the raw sequence read counts. A heatmap of the results is given in Figure 2. Twenty-one probes were found to be positive relative to the negative controls. The positive probes correspond to seventeen viruses (Figure 2). Surprisingly, more viruses were found in the healthy controls than in the ME/CFS patients. No virus was found to be unique to the patients. No virus was found systematically in all ME/CFS samples. However, a probe mapping to human alphaherpesvirus 3 (Probe ID: NC_001348_4033) showed the strongest signal, with both the largest effect size and the most significant p-value (Table 1, Figure 2). As seen in Figure 2, this probe is elevated in six ME/CFS patients and absent in all healthy controls.

**Figure 2.**
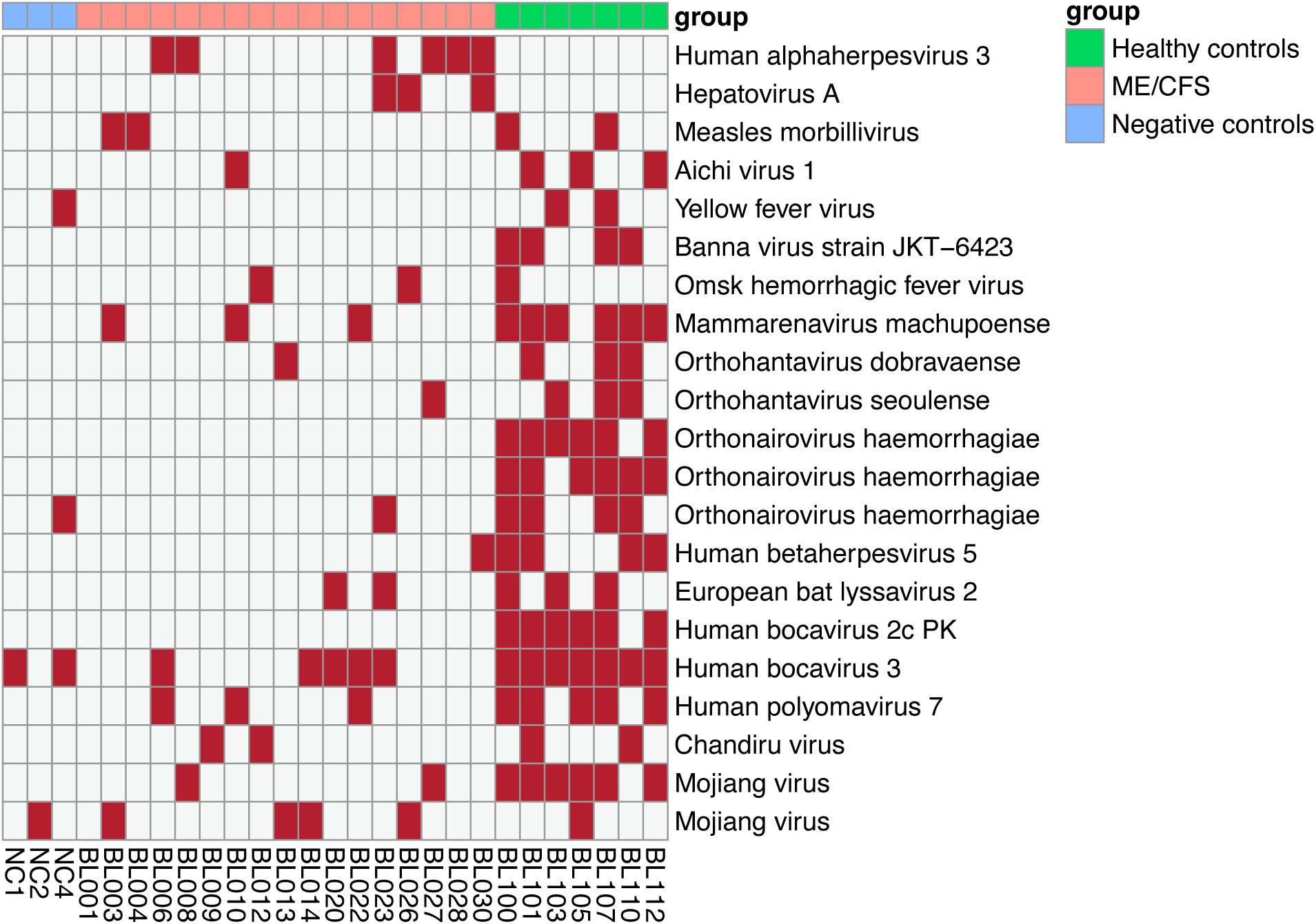
Virus sequences detected in each clinical sample. This figure presents a heatmap of those molecular probes which are significantly enriched. The rows correspond to individual probes. The right labels present the virus name. The columns correspond to the individual clinical samples. The top color bar indicates the group (blue, negative controls [water reacted with the probe ensemble; no DNA present]; pink, ME/CFS patients; green, healthy controls.) The bottom labels give the identification of each clinical sample. Cell colors indicate the presence or absence of the molecular probe in the sample (red, presence; white, absence).

## Discussion

Viruses are often found in the blood of hemorrhagic fever patients (Kallio-Kokko et al. 2005): e.g., Dengue virus (Martina 2014) and Hantaviruses (Noack et al. 2020). However, these patients are not of concern here. Rather, we are interested in the viruses in the blood of healthy people (reviewed in Rascovan et al. 2016) and ME/CFS patients. As one example, Moustafa et al. identified 19 human viruses in the blood of healthy individuals (Moustafa et al. 2017). Human herpesvirus 4 (HHV-4, formerly known as Epstein-Barr virus) was found in 14.45% of individuals, and Hepatitis C virus was identified in 0.01% of individuals. HHV-7 was found most frequently (20.37%). Our molecular probes for HHV-7 did not detect the genome of this virus in our clinical samples. As a second example, Furuta et al. (2015) employed metagenomic profiling and PCR to identify viruses in the blood of blood donors in Japan. Two human herpesviruses were detected: HHV-4 and HHV-6B, as were mastadenovirus, Torque-Teno Virus, picornaviruses, and flaviviruses. Thus, the viral genome sequences that we identified in the cell-free blood of healthy controls are consistent with those identified by previous authors (c.f., Haynes and Rohwer 2010; Liang and Bushman 2021).

The molecular probes contain 60 bases of genomic DNA. Even in those cases where three probes were positive for a given virus, at this low coverage, it cannot be determined if the entire genome sequence was present. Therefore, we consider our virus identifications as preliminary. They should be confirmed by further experiments.

## Data Availability

All scripts, keyfiles, and data (starting with the collapsed fastq sequences at the end of step2) are available at https://github.com/k-roy/MOLECULAR_PROBES and archived with Zenodo at https://doi.org/10.5281/zenodo.17429326.

https://doi.org/10.5281/zenodo.17429326

## Supplementary information

**Figure S1.**
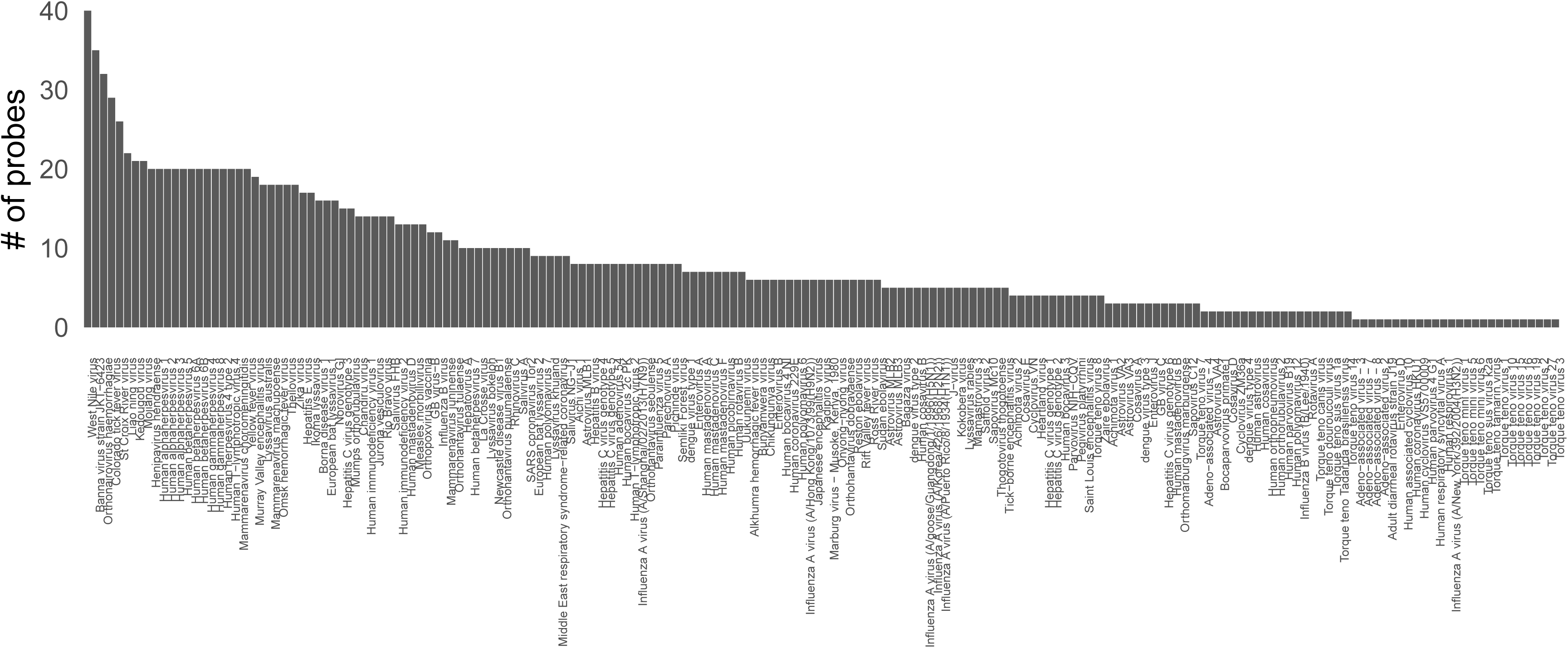
Molecular probe ensemble. The distribution of the number of probes per virus applied in this study.

**Figure S2.**
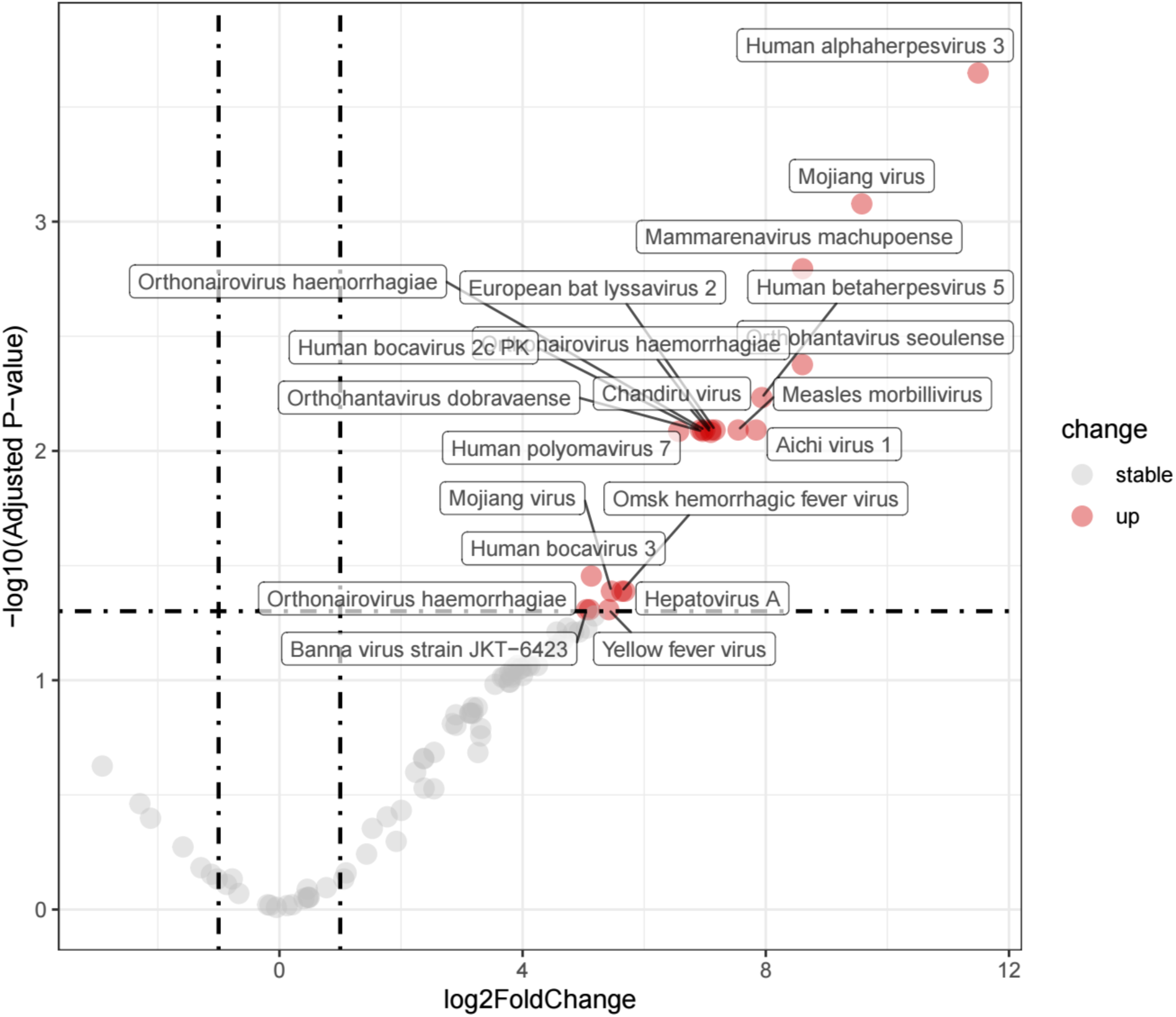
Volcano plot of differential analysis of molecular probes. Each point is a probe; red points are molecular probes significantly enriched in CFS patients versus negative controls (FDR < 0.05 and |log2 fold change|>1). Labels indicate the corresponding virus (multiple probes may map to the same virus). The x-axis shows log2 fold change (CFS vs. control); the y-axis shows −log10 of the Benjamini–Hochberg–adjusted p-value (FDR). Vertical dashed lines mark |log2 fold change| = 1 (i.e., a 2-fold change), and the horizontal dashed line marks FDR = 0.05.

**Table S1.**
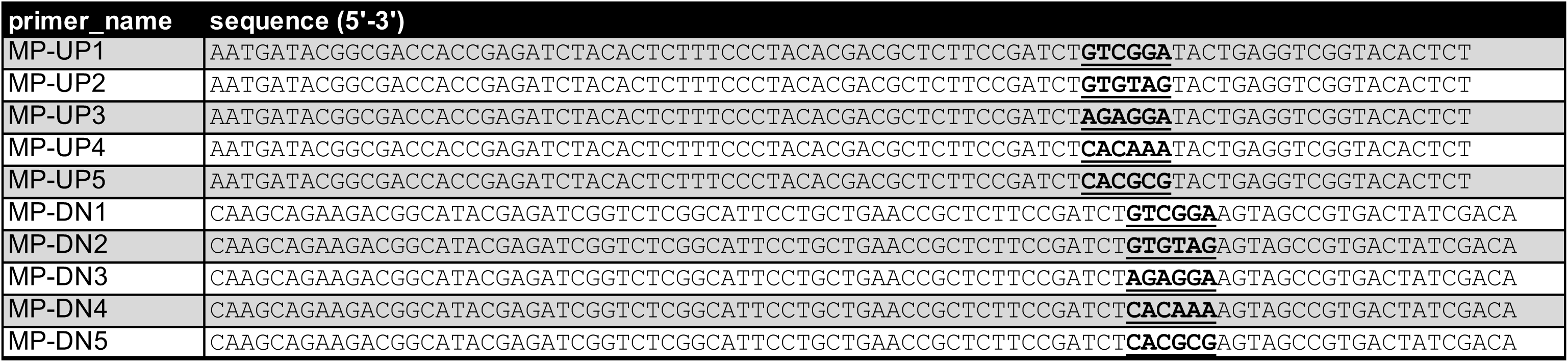
Sequencing primers.

**Table S2.** Primer mixes.

| primer_mix | R1_bc_name | R2_bc_name |
| --- | --- | --- |
| A | MP-UP1 | MP-DN1 |
| B | MP-UP1 | MP-DN2 |
| C | MP-UP1 | MP-DN3 |
| D | MP-UP2 | MP-DN1 |
| E | MP-UP2 | MP-DN2 |
| F | MP-UP2 | MP-DN3 |
| G | MP-UP3 | MP-DN1 |
| H | MP-UP3 | MP-DN2 |
| I | MP-UP3 | MP-DN3 |
| J | MP-UP1 | MP-DN4 |
| K | MP-UP1 | MP-DN5 |
| L | MP-UP2 | MP-DN4 |
| M | MP-UP2 | MP-DN5 |
| N | MP-UP3 | MP-DN4 |
| O | MP-UP3 | MP-DN5 |
| P | MP-UP4 | MP-DN1 |
| Q | MP-UP4 | MP-DN2 |
| R | MP-UP4 | MP-DN3 |
| S | MP-UP4 | MP-DN4 |
| T | MP-UP4 | MP-DN5 |
| U | MP-UP5 | MP-DN1 |
| V | MP-UP5 | MP-DN2 |
| W | MP-UP5 | MP-DN3 |
| X | MP-UP5 | MP-DN4 |
| Y | MP-UP5 | MP-DN5 |

**Table S3.**
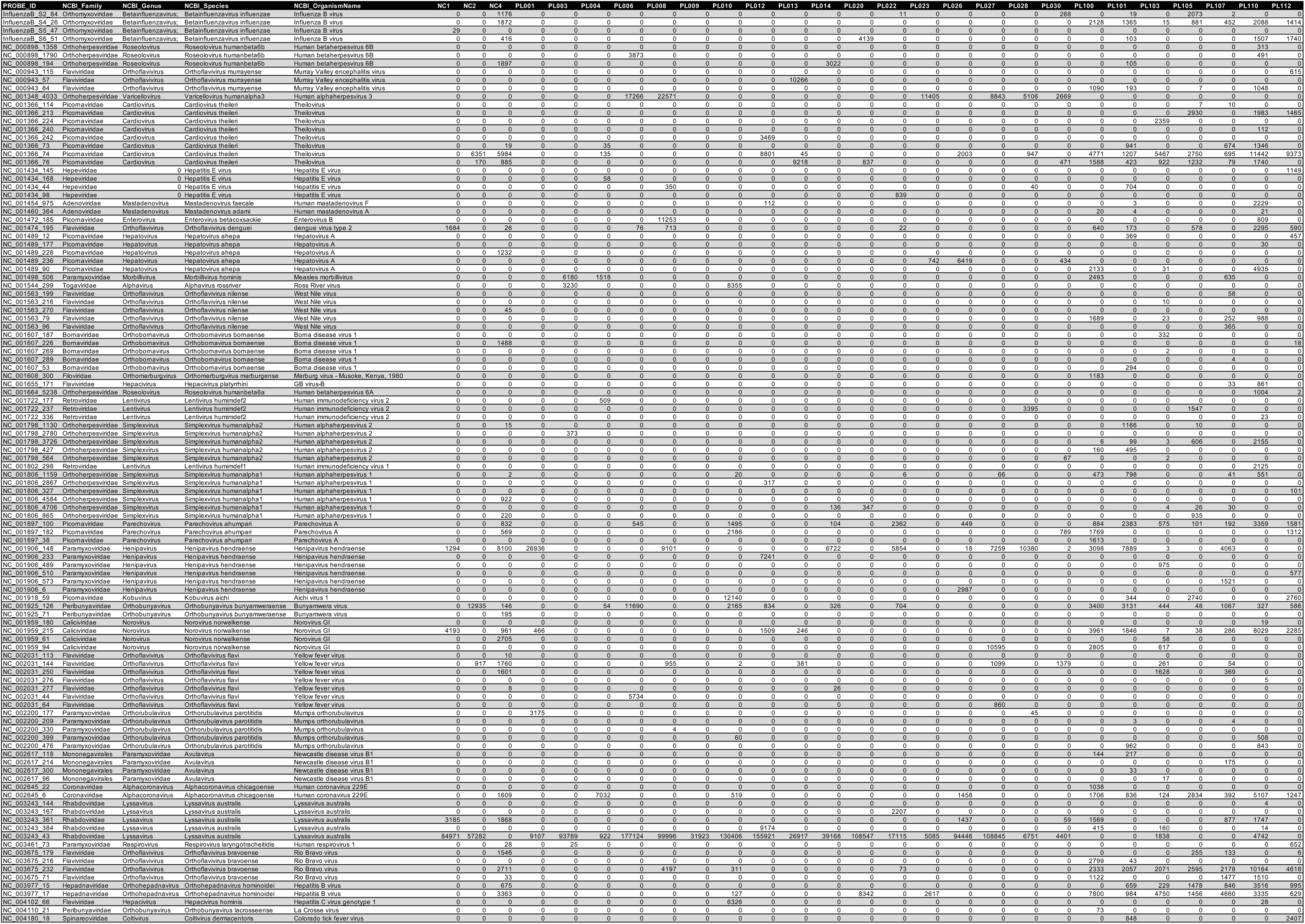
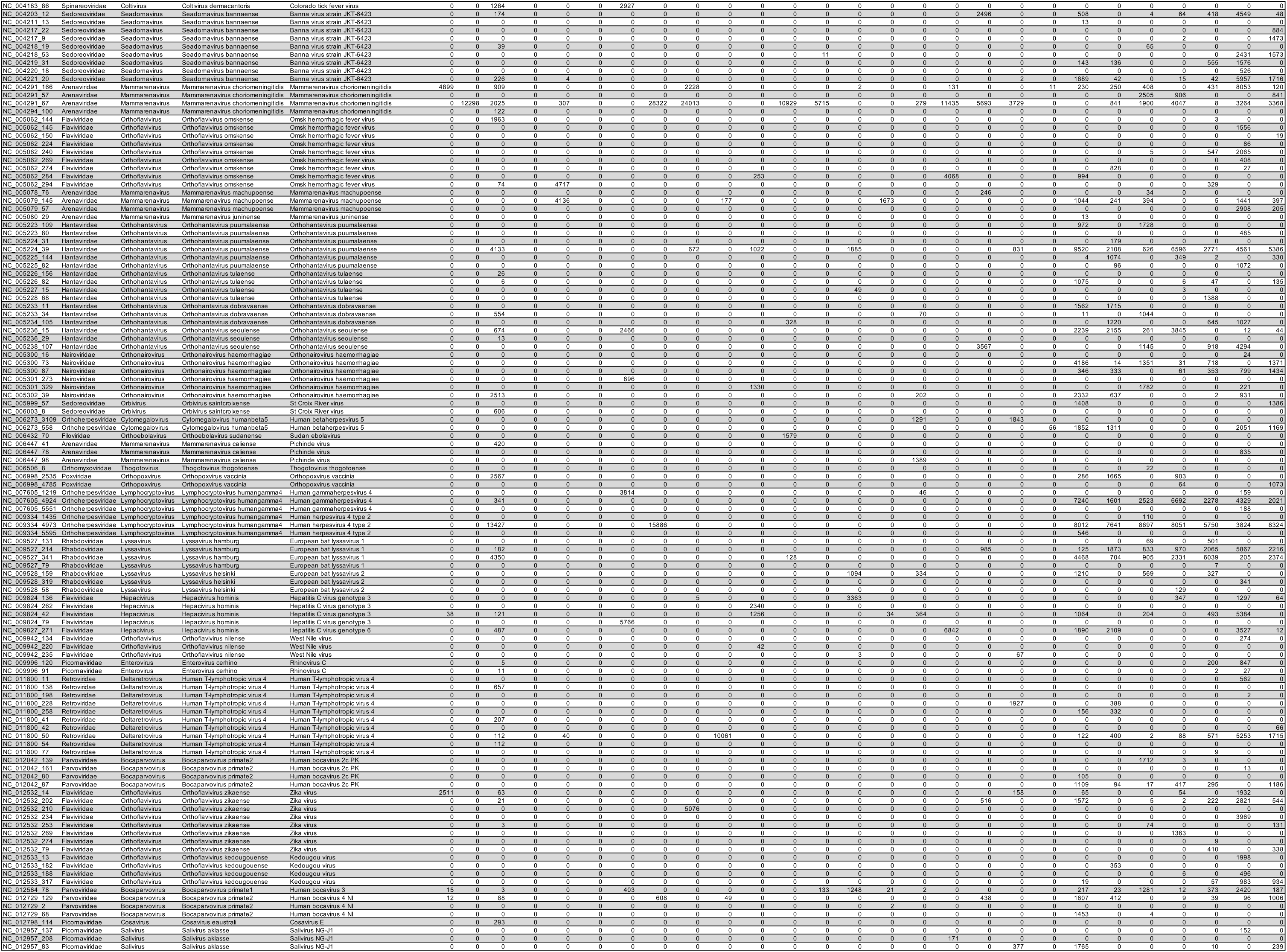
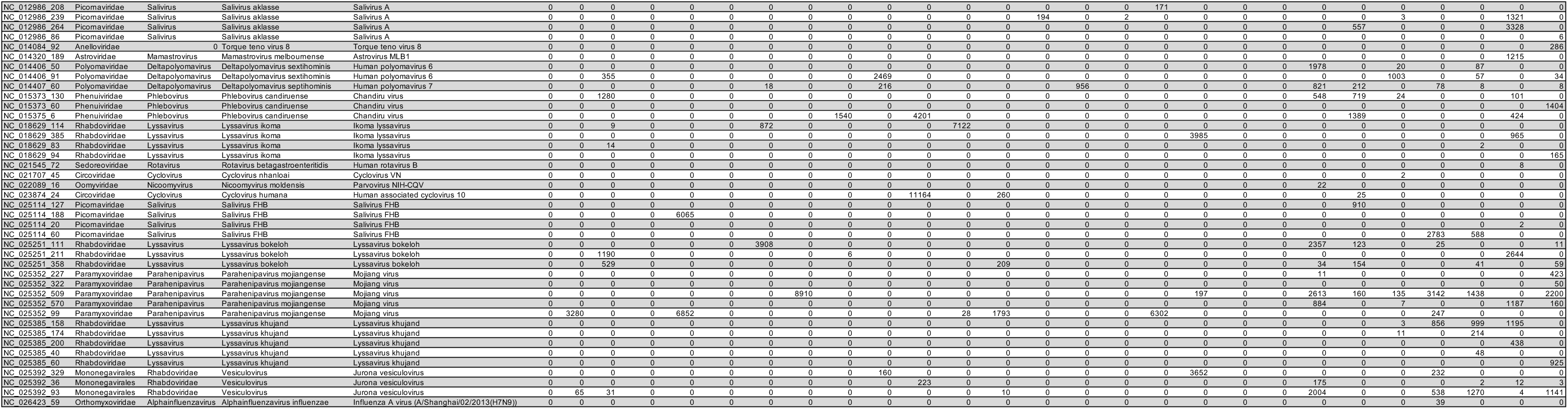
Read counts.

